# RAX-NET: Residual Attention Xception Network for Brain Ischemic Stroke Segmentation in T1-Weighted MRI

**DOI:** 10.1101/2025.09.26.25336733

**Authors:** Amir Mousavi

## Abstract

Ischemic stroke, caused by arterial occlusion, leads to hypoxia and cellular necrosis. Rapid and accurate delineation of ischemic lesions is essential for treatment planning but remains challenging due to variations in lesion size, shape, and appearance. We propose Residual Attention Xception Network, a deep learning architecture that integrates residual attention connections with Xception for three-dimensional magnetic resonance imaging lesion segmentation. The framework includes three stages: (i) decomposition of three-dimensional scans into axial, sagittal, and coronal planes, (ii) independent model training on each plane, and (iii) voxel-wise majority voting to generate the final three-dimensional segmentation. In addition, we introduce a variant of the focal Tversky loss designed to mitigate class imbalance and improve sensitivity to small or irregular lesion boundaries. Experiments on the ATLAS v2.0 dataset with five-fold cross-validation demonstrate that Residual Attention Xception Network achieves a Dice coefficient of 0.61, precision of 0.68, and recall of 0.63. These results surpass baseline models while requiring fewer trainable parameters and enabling faster inference, highlighting both accuracy and efficiency.

**source code:** https://github.com/liamirpy/RAX-NET_ISCHEMIC_STROKE_SEGMENTATION.

## I. Introduction

**S**TROKE is a leading cause of disability and mortality worldwide [1]. It is broadly classified into ischemic, caused by infarction, and hemorrhagic, caused by bleeding [2]. Ischemic stroke predominates, accounting for approximately 62% of cases globally and 87% in the United States [1]. Classification of ischemic stroke depends on the time since symptom onset, with acute, subacute, and chronic phases corresponding to within the first day, one to two weeks, and more than two weeks post-onset, respectively [3]. Lesion–symptom mapping, which examines the relationship between lesion location and clinical symptoms, relies heavily on precise segmentation [4]. Similarly, in clinical settings, rapid and accurate delineation of ischemic lesions is essential for treatment planning. However, segmentation remains challenging due to variability in lesion size, shape, and intensity, as well as limited availability of multimodal imaging. MRI modalities are generally favored for their higher sensitivity and superior contrast compared to CT in stroke lesion detection [5], [6].While multimodal MRI can improve segmentation performance, acquisition of such datasets is often impractical in clinical and research contexts due to cost, availability, and patient-related constraints [7]. To address this limitation, researchers frequently resort to single-modality approaches, with a preference for T1-weighted (T1w) images. T1w MRI, known for its excellent spatial resolution and relatively short acquisition duration, is also commonly used to register other imaging modalities, such as functional MRI and diffusion MRI.

Manual segmentation, although considered the benchmark for stroke lesion delineation, is labor-intensive and prone to inter- and intra-observer variability. Despite the development of various tools for lesion segmentation in T1w images, accuracy often depends on lesion size and location. The challenge is further exacerbated by the similarity in gray levels between lesions and adjacent regions, particularly the cerebrospinal fluid (CSF), coupled with variations in lesion shape. Consequently, substantial manual refinement is often required [8]. Moreover, segmentation performance is influenced by the stroke phase (acute, subacute, or chronic) at the time of imaging.

In recent years, deep learning methods have shown promise in medical image analysis, particularly for stroke lesion segmentation [9]–[11]. These approaches have the potential to overcome limitations of manual segmentation by providing more efficient and accurate solutions for lesion identification and delineation in T1w MRI.

This work makes three primary contributions. First, it provides a systematic review of existing lesion segmentation approaches and publicly available datasets across the acute, subacute, and chronic phases of ischemic stroke. Second, it introduces Residual Attention Xception Network (RAX-Net), a deep learning framework that integrates residual attention mechanisms with the Xception backbone to enable accurate and efficient segmentation of ischemic stroke lesions in T1w MRI. Third, it presents a comprehensive evaluation of RAX-Net on the Anatomical Tracings of Lesions After Stroke (ATLAS) v2.0 dataset [12], which contains high-quality, expert-annotated T1w images. The remainder of this paper is organized as follows. Section II reviews existing segmentation methods and datasets. Section III describes the proposed RAX-Net architecture. Section IV outlines the experimental setup and evaluation protocols. Section V reports the results and comparative analyses. Finally, Section VII summarizes the findings and discusses future directions.

## II. RELATED WORK

### A. Acute: ISLES2015-SPES

The ISLES2015-SPES dataset [3] comprises 50 patient cases, with lesion volumes ranging from 7.3 × 10^3^ mm^3^ to 361.8 × 10^3^ mm^3^. According to the review in [13], models trained on this dataset achieved Dice coefficients between 0.79 and 0.88. Among the most widely adopted architectures, U-Net [14], originally proposed for biomedical image segmentation, has demonstrated consistent results. Li et al. [15] proposed a multi-kernel DCNN based on U-Net, consisting of two parallel U-Net branches trained on multimodal MRI data with preprocessing and postprocessing. On the ISLES2015-SPES test set, the model achieved a Dice score of 0.79, though all images were resized to 160 × 160, which may limit generalizability. Sathish et al. [16] employed multisequence MRI with cross-entropy loss and three discriminators using relativistic visual Turing tests, achieving a Dice coefficient of 0.82 with three-fold cross-validation. Only Dice, recall, and precision were reported. Kuma et al. [17] proposed the Classifier-Segmenter Network (CSNet), combining a 2D classifier with a 3D segmenter. Using five-fold cross-validation, the model achieved a Dice coefficient of 0.83, though classifier evaluation was not reported. Albert et al. [18] developed an asymmetrical residual 3D CNN that reached a Dice of 0.84, but required high computational resources. Rajan et al. [19] introduced the SUMNet architecture, reporting a Dice of 0.88; only Dice was reported. Two additional acute-phase datasets are available: ISLES2017 and ISLES2018, with Dice coefficients reported in the ranges of [0.23, 0.34] and [0.49, 0.51], respectively [13].

### B. Subacute: ISLES2015-SISS

The ISLES2015-SISS dataset [3] contains 64 patient cases with lesion volumes between 1.0 × 10^3^ mm^3^ and 346.1 × 10^3^ mm^3^. Reported Dice coefficients range from 0.55 to 0.76. Kamnitsas et al. [20] introduced a 3D CNN with architectural modifications to reduce complexity and training time, achieving a Dice of 0.59 on 90 test MRI scans. Zhang et al. [21] proposed the multi-plane fusion network (MPFN), which detects regions of interest (ROI) in axial, sagittal, and coronal planes, followed by ROI segmentation via a fusion network. The model achieved a Dice of 0.62. CSNet [17] achieved a Dice of 0.63, while Liu et al. [22] introduced DRANet, incorporating attention mechanisms to improve small-lesion detection. By decomposing 3D MRI into 2D slices, DRANet achieved a Dice of 0.76, although only 224 2D test images were used.

### C. Chronic: ATLAS v1.2

The ATLAS v1.2 dataset [12] consists of 304 cases, with lesion volumes ranging from 0.01 × 10^3^ mm^3^ to 283.8 × 10^3^ mm^3^. One study [23] proposed dimension-fusion U-Net (D-U-Net), which combines 2D and 3D models. On a 20% test split, the method achieved a Dice of 0.53, reporting only Dice, recall, and precision, and lacking comparisons to ischemic stroke–specific methods. Another work proposed MSDF-Net with atrous spatial pyramid pooling (ASPP) [24], achieving a Dice of 0.56 on 69 validation cases, reporting Dice, sensitivity, and volumetric overlap error. Qi et al. [25] introduced X-Net, which incorporates a feature similarity module (FSM) to capture long-range dependencies. Using five-fold cross-validation, X-Net achieved a Dice of 0.48. Yang et al. [26] proposed CLCI-Net, combining cross-level feature fusion and ConvLSTM for contextual modeling, achieving a Dice of 0.58 on 60 test cases. Hui et al. [27] developed partitioning-stacking prediction fusion (PSPF), based on attention U-Net. The model fused 2D predictions from different planes into a 3D mask, achieving a Dice of 0.59 with six-fold cross-validation. Tomita et al. [28] proposed a modified 3D U-Net with a zoom in/out strategy, achieving an average Dice of 0.64 on 31 test cases and 0.80 for large lesions, but only 0.41 for small lesions. Training required substantial GPU resources.

### D. Acute, Subacute, Chronic: ATLAS v2.0

The v2.0 dataset contains 955 cases, of which 655 include expert-annotated lesion masks. Each case has a single 3D lesion mask reviewed by two expert teams, with rigorous quality control. Scans were acquired using 1.5T and 3T MRI scanners, with voxel sizes of ≤ 1 mm^3^ in most cases. Four subsets include 1–2 mm^3^ resolution in at least one dimension. All images are single-modality T1-weighted MRI.

#### 1) Lesion Distribution Pattern

Metadata for each lesion mask includes lesion count, lateralization (left or right hemisphere), and anatomical categorization (cortical or subcortical).

#### 2) Hemisphere Location

Among the 655 cases, 61.9% contain a single lesion, while 38.1% have multiple lesions. For single-lesion cases, 26.4% are in the left hemisphere, 28.5% in the right, and 7.0% in other regions. For multiple-lesion cases, 7.2% are unilateral, 18.5% bilateral, and 12.4% in other locations, such as the cerebellum or brainstem (Table I).

**TABLE I.** LESION COUNT AND LOCATION IN CORTICAL AND SUBCORTICAL REGIONS OF THE ATLAS V2.0 DATASET.

|  | One Lesion |  |  | Multiple Lesions |  |  |
| --- | --- | --- | --- | --- | --- | --- |
|  | Left | Right | Other | Unilateral | Bilateral | Other |
| Data | 173 (26.4%) | 187 (28.5%) | 46 (7.0%) | 47 (7.2%) | 121 (18.5%) | 81 (12.4%) |

#### 3) Subcortical vs. Cortical Location

Of the 1101 lesions, 25.5% are cortical, 59.6% subcortical, and 14.8% in other regions. Distribution across hemispheres is detailed in Table II and Fig. 1.

**TABLE II.** LESION COUNT AND LOCATION IN CORTICAL AND SUBCORTICAL REGIONS OF THE ATLAS V2.0 DATASET.

|  | One Lesion |  | Multiple Lesions |  | Others | Total |
| --- | --- | --- | --- | --- | --- | --- |
|  | Left | Right | Left | Right |  |  |
| Data | 132 (12.0%) | 149 (13.5%) | 333 (30.2%) | 324 (29.4%) | 163 (14.8%) | 1101 |

**Fig. 1.**
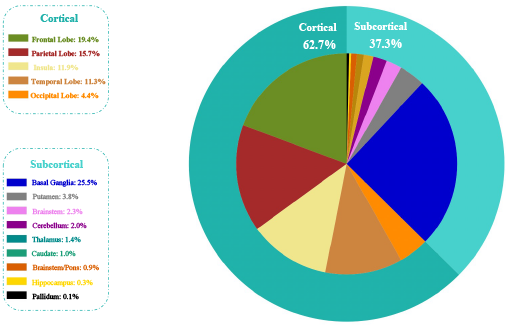
Distribution of ischemic stroke lesions across cortical and subcortical regions in the ATLAS v2.0 dataset.

### E. Loss Functions

Loss functions are central to segmentation performance. In binary segmentation, binary cross-entropy [29] is widely used to measure pixel-wise prediction error. For overlap-based optimization, the Dice loss [30] maximizes region similarity. Tversky loss [31], a generalization of Dice, addresses class imbalance by weighting false positives and false negatives. Focal loss [32] further mitigates imbalance by emphasizing hard-to-classify samples.

## III. METHOD

The proposed methodology consists of three main components: image plane decomposition, the segmentation model, and an aggregation function. The first subsection describes the decomposition of 3D MRI volumes into 2D slices. The second subsection details the segmentation framework. The final subsection introduces the aggregation function, which fuses predictions across planes to produce the final 3D segmentation.

### A. IMAGE PLANE DECOMPOSITION

Brain magnetic resonance imaging volumes are three-dimensional tensors that can be viewed along three orthogonal planes: axial, sagittal, and coronal. Each plane provides complementary anatomical perspectives. The axial plane divides the brain into superior and inferior sections, with the x-axis representing left–right and the y-axis representing anterior–posterior directions. The sagittal plane partitions the brain into left and right halves, with the y-axis representing anterior–posterior and the z-axis representing superior–inferior directions. The coronal plane separates anterior and posterior sections, with the x-axis representing left–right and the z-axis representing superior–inferior directions.

Both 3D and 2D segmentation models have been widely explored. While 3D models capture volumetric context effectively, they require substantial computational resources. In contrast, 2D models trained on single planes are more efficient but risk losing cross-plane contextual information. Importantly, lesions that appear ambiguous in one plane may be more clearly delineated in another. To address this, all three planes are incorporated into the segmentation pipeline, with predictions combined through an aggregation function to improve accuracy. Accordingly, in the first stage of our approach, MRI volumes are decomposed into 2D slices along the axial, sagittal, and coronal planes (Fig. 2).

**Fig. 2.**
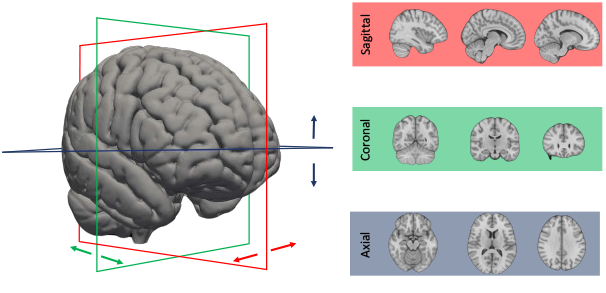
Decomposition of a 3D MRI scan into axial, sagittal, and coronal planes. Blue, red, and green rectangles indicate the superior–inferior (axial), anterior–posterior (sagittal), and left–right (coronal) axes, respectively.

### B. RAX-NET

The proposed RAX-Net architecture integrates convolution, separable convolution, up-sampling, and transposed convolution layers. Residual connections are included in the encoder to mitigate feature loss during down-sampling, while an up-sampling residual attention block enhances accuracy in the decoder. The network follows a U-shaped design with:

#### Encoder (contracting path)

convolutional and separable convolutional blocks progressively extract deeper features while reducing spatial resolution. Residual connections preserve fine-grained details.

#### Decoder (expanding path)

transposed convolutional layers restore spatial resolution, and residual attention blocks fuse contextual and fine-grained features. This design captures both local and global context for accurate lesion segmentation. The proposed architecture represented in Figure 4.

#### 1) Separable Convolution Layer

In conventional neural networks (CNNs), each convolutional filter operates simultaneously across both the spatial dimensions and the channel dimension of the input feature maps. While this approach provides strong representational power, it is computationally expensive and parameter-heavy, especially for high-resolution medical images. To address this issue, the proposed architecture incorporates *separable convolution*, which factorizes a standard convolution into two sequential operations: *depthwise convolution* and *pointwise convolution*. This decomposition reduces computational complexity while maintaining feature extraction capabilities. An illustration is shown in Fig. 3.

**Fig. 3.**
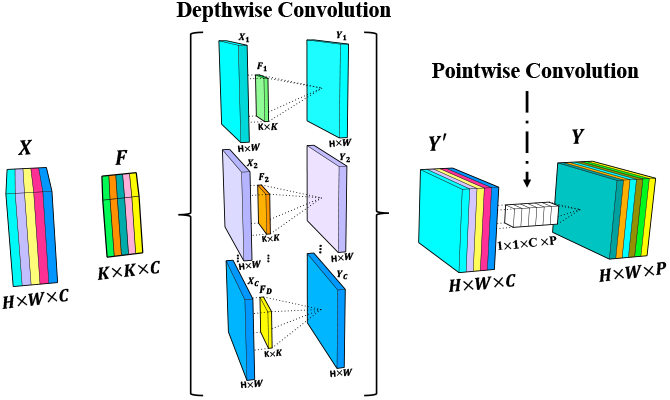
Illustration of the separable convolution architecture, showing the decomposition of standard convolution into depthwise and pointwise operations.

**Fig. 4.**
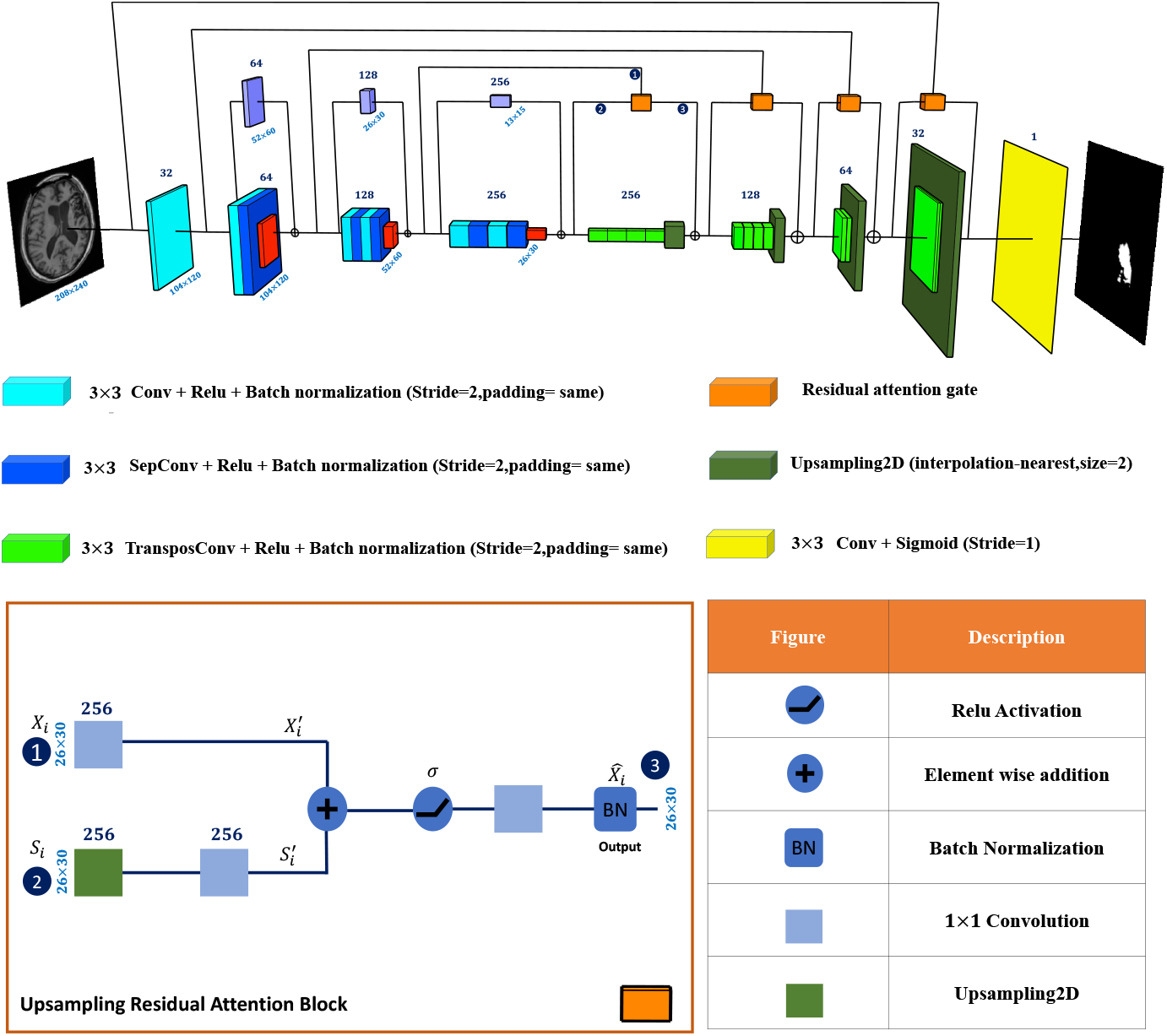
Architecture of the proposed RAX-Net, showing encoder–decoder design, residual connections, and residual attention blocks.

#### Depthwise convolution

Given an input tensor 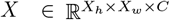, where *X*_*h*_, *X*_*w*_, and *C* denote height, width, and number of channels, respectively, depthwise convolution applies a distinct 2D filter to each channel independently. Let 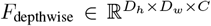 be the set of depthwise filters, where *D*_*h*_ and *D*_*w*_ are the spatial dimensions of the kernel. The output of the depthwise convolution, denoted by *Y* ^*′*^, is computed as:

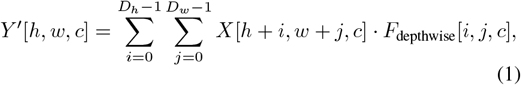

for each spatial location (*h, w*) and channel *c*. This operation captures local spatial correlations within each individual channel.

#### Pointwise convolution

Following depthwise convolution, a 1 × 1 convolution, referred to as pointwise convolution, is applied to combine information across channels. Let *F*_pointwise_ ∈ ℝ^1*×*1*×C×P*^ denote the pointwise filter, where *P* is the number of output channels. The final output tensor 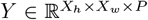 is computed as:

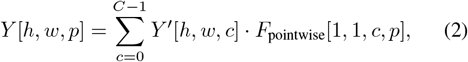

for each output channel *p*. This operation integrates information across the channels processed by the depthwise convolution.

#### Advantages

The separable convolution layer reduces both the number of parameters and the computational load compared to traditional convolution. This efficiency is particularly beneficial for large-scale 3D medical imaging tasks. Moreover, depthwise convolution captures fine-grained spatial patterns within each channel, while pointwise convolution aggregates cross-channel information, effectively balancing local detail extraction with global feature integration. However, since depthwise and pointwise operations are decoupled, the expressive capacity may be slightly reduced compared to full convolutions.

#### 2) Up-Sampling Residual Attention Block

The up-sampling residual attention block is designed to combine spatial detail from encoder features with contextual information from decoder features, thereby enhancing segmentation of lesions with complex boundaries.

Let the block receive two inputs:

- *X*_*i*_: feature maps from the encoder (down-sampling path), rich in spatial detail but limited in semantic context.
- *S*_*i*_: feature maps from the decoder (up-sampling path), rich in semantic information but with lower spatial resolution.

First, the decoder feature *S*_*i*_ is up-sampled to match the spatial resolution of *X*_*i*_. Then, both *S*_*i*_ and *X*_*i*_ are projected via 1 × 1 convolutions into aligned feature spaces, producing 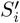 and 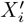. These two representations are fused through element-wise addition:

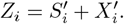

The combined features *Z*_*i*_ are passed through a nonlinear activation (ReLU), followed by a convolution and batch normalization layer:

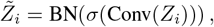

where *σ*(·) denotes the ReLU activation function and BN denotes batch normalization.

Finally, a residual connection ensures that fine-grained encoder features are preserved alongside the refined output. This mechanism stabilizes learning and improves boundary delineation for lesions with irregular shapes or low contrast.

### C. Aggregation Function

After independent segmentation in each imaging plane, predictions are aggregated to generate the final 3D mask.

Given an MRI volume *M* ∈ ℝ^197*×*233*×*189^, we extract three sets of 2D slices:

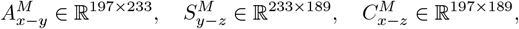

corresponding to axial, sagittal, and coronal planes, respectively.

Plane-specific models *f*_*x*−*y*_, *f*_*y*−*z*_, *f*_*x*−*z*_ predict lesion probability maps:

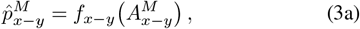

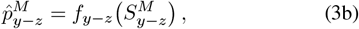

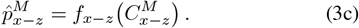

The aggregated probability map is computed by averaging the three predictions:

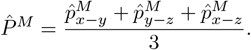

A thresholding operator *T* (·) with *τ* = 0.5 is then applied to produce the final binary mask:

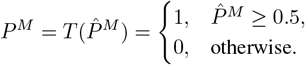

#### Interpretation

This aggregation ensures that voxel-level predictions are confirmed by the majority of plane-specific models, thereby reducing false positives from any single view. Thresholding at 0.5 corresponds to a simple majority vote. The procedure leverages complementary information from axial, sagittal, and coronal planes, enabling robust 3D segmentation even when lesion boundaries are ambiguous in individual planes (Fig. 5).

**Fig. 5.**
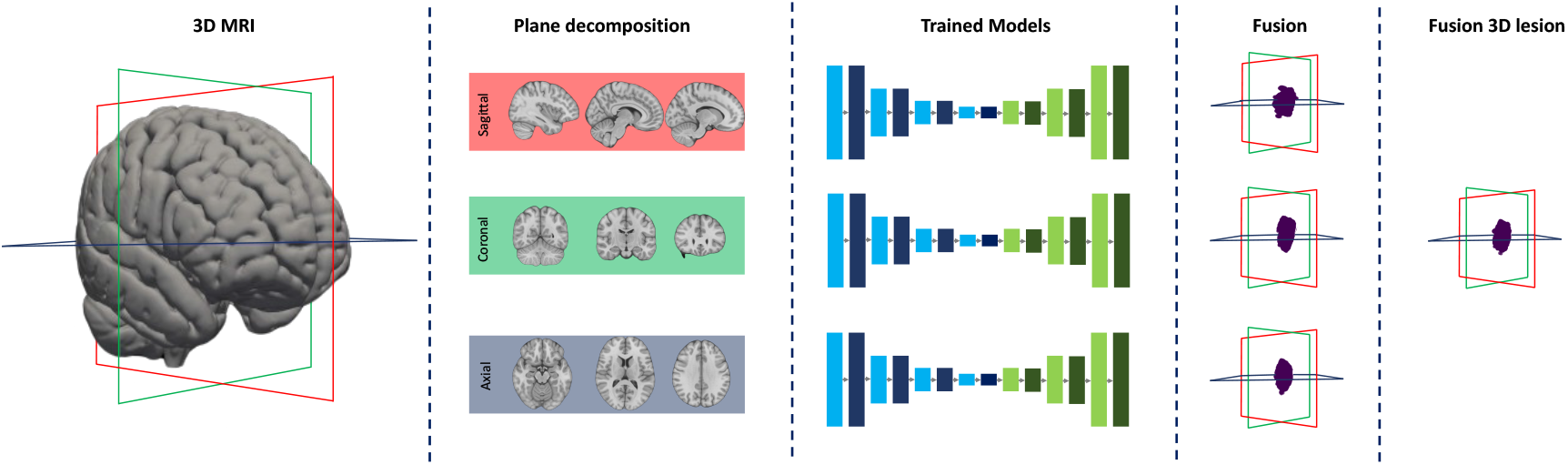
Workflow of the proposed approach: (i) decomposition of 3D MRI into 2D slices, (ii) per-plane lesion segmentation using 2D models, and (iii) aggregation of predictions into a final 3D mask.

## IV. Experiments

### A. Data Division

This study utilizes the ATLAS v2.0 database, focusing on the 655 cases with expert-annotated T1-weighted 3D lesion masks. The dataset was divided into two disjoint subsets: (i) a set for training and evaluating the plane-specific 2D models and (ii) a held-out set for evaluating the aggregation procedure and the final 3D segmentation. Nineteen cases were excluded due to insufficient image quality, resulting in 636 usable cases. Of these, 524 cases were used for training and evaluation of the 2D models, while 112 cases were reserved for assessment of the aggregation function. The lesion voxel-size distributions for the two subsets are shown in Fig. 6(a).

**Fig. 6.**
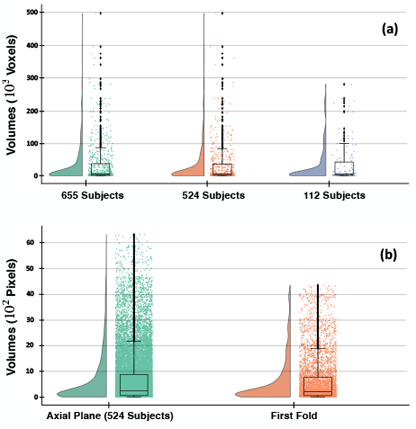
Lesion distributions in ATLAS v2.0. (a) Lesion voxel-size distribution for the 524 cases used to train/evaluate 2D models and the 112 cases reserved for aggregation and 3D segmentation. (b) Axial-plane lesion pixel-size distribution for the 524-case set and for one representative cross-validation fold.

### B. Cross-Validation

For training the 2D models, each of the 524 cases was decomposed into axial, sagittal, and coronal slice stacks. Three plane-specific models were trained independently using five-fold cross-validation. To stratify subjects according to lesion size, predefined intervals were employed:

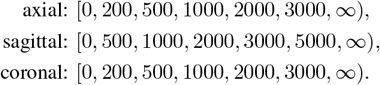

Folds were generated such that (i) the lesion-size distribution in each fold approximated that of the complete dataset for the corresponding plane and (ii) all slices from a given subject were assigned to the same fold to prevent information leakage. Each training fold contained an equal number of lesion and non-lesion slices, yielding balanced training sets and ensuring robust evaluation. An example of the axial-plane lesion-size distribution for the complete dataset and for one fold is shown in Fig. 6(b).

### C. Distribution-Aware Batching

During training, mini-batches were constructed in a distribution-aware manner to mitigate class imbalance and account for heterogeneity in lesion size. Prior to sampling, lesion slices were grouped into bins according to lesion size, and non-lesion slices were indexed separately. Each minibatch was then formed by sampling lesion slices proportionally from each bin and pairing them with an equal number of non-lesion slices. This strategy preserved the global lesion-size distribution across training batches while ensuring balanced class representation. Owing to the diverse sources of ATLAS data, the risk of overfitting under this approach was reduced. Furthermore, Batch Normalization benefited from more stable within-batch statistics, contributing to accelerated convergence.

### D. Evaluation Metrics

Performance was assessed using precision, recall, and the Dice similarity coefficient (DSC), computed from true positives (TP), false positives (FP), false negatives (FN), and true negatives (TN). Let *y* ∈ {0, 1}^|Ω|^ denote the ground-truth mask and *ŷ* ∈ {0, 1} ^|Ω|^ the predicted mask over voxel set Ω. Then,

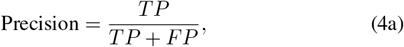

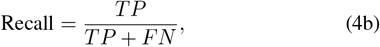

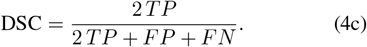

Precision quantifies reliability of positive predictions, recall measures sensitivity to true lesions, and DSC evaluates spatial overlap between predictions and reference annotations.

### E. Loss Functions

Let *p*_*v*_ ∈ [0, 1] denote the predicted probability of voxel *v* ∈ Ω being lesion and *g*_*v*_ ∈ {0, 1} its ground-truth label. The following loss functions were employed.

#### 1) Binary Focal Cross-Entropy

The focal loss modifies binary cross-entropy by down-weighting easy examples and focusing on hard examples:

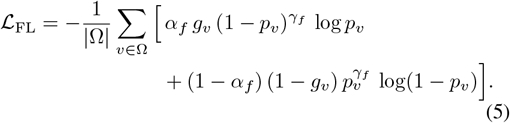

where *α*_*f*_ balances lesion and background contributions and *γ*_*f*_ controls the focusing strength.

#### 2) Tversky Loss

The Tversky index generalizes Dice by asymmetrically penalizing false positives and false negatives:

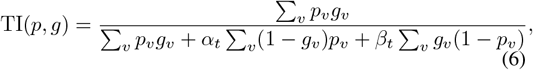

with *α*_*t*_ and *β*_*t*_ weighting FP and FN, respectively. The Tversky loss is then

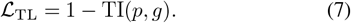

#### 3) Focal Tversky Loss (Proposed Variant)

To simultaneously address class imbalance and emphasize difficult examples, a convex combination of focal loss and Tversky loss was employed:

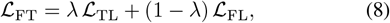

where *λ* ∈ [0, 1] is a mixing coefficient. When *λ* is closer to 1, training emphasizes overlap via Tversky loss, whereas smaller *λ* values increase the contribution of hard-example mining from focal loss. This formulation improves boundary delineation and robustness to lesions of varying size and contrast.

### F) Training Protocol

All models were implemented in Keras/TensorFlow and trained on an NVIDIA GeForce GTX 1080-Ti GPU. Optimization was performed using AdamW with an initial learning rate of 0.01 and exponential decay. Mini-batches contained 32 slices (16 lesion and 16 non-lesion). Early stopping based on validation loss was applied to prevent overfitting. Input dimensions were zero-padded to (208 × 240) for axial, (240 × 208) for sagittal, and (208 × 208) for coronal slices. Unless otherwise noted, loss hyperparameters were set to *α*_*f*_ = 0.8, *γ*_*f*_ = 2.0, (*α*_*t*_, *β*_*t*_) = (0.3, 0.7), and *λ* tuned based on validation performance.

## V. Results

A comprehensive evaluation was conducted to assess each component of the proposed framework. Results are organized into three parts: (i) loss-function analysis, (ii) segmentation model evaluation and comparison, and (iii) aggregation-function evaluation.

### A. Loss-Function Analysis

To examine the effect of the loss function on segmentation performance, the first axial fold was trained using four losses: Dice loss, focal loss, Tversky loss, and the proposed focal–Tversky (FocalTversky) loss. The results in Table III indicate that FocalTversky achieved the highest Dice and F1, with a balanced trade-off between precision and recall. Dice loss yielded the highest precision, whereas Tversky loss produced the highest recall.

**TABLE III.** Performance of Different Loss Functions on the First Axial Fold.

| Loss Function | Dice | Precision | Recall | F1 |
| --- | --- | --- | --- | --- |
| Dice Loss | 0.7859 | **0.8430** | 0.7406 | 0.7889 |
| Focal Loss | 0.7326 | 0.7302 | 0.7427 | 0.7364 |
| Tversky Loss | 0.7764 | 0.7559 | **0.8037** | 0.7791 |
| FocalTversky (Proposed) | **0.7904** | 0.7963 | 0.7894 | **0.7928** |

Note: For binary segmentation, Dice and F1 are numerically equivalent under the same TP/FP/FN counts; both are reported for completeness.

### B. Segmentation Model Evaluation

#### 1) Proposed Model: Cross-Validation Performance

Five-fold cross-validation was performed independently per plane. Table IV summarizes the per-fold performance and the across-fold mean ± standard deviation. The axial and coronal models obtained higher Dice and precision on average, while recall remained competitive across planes.

**TABLE IV.**
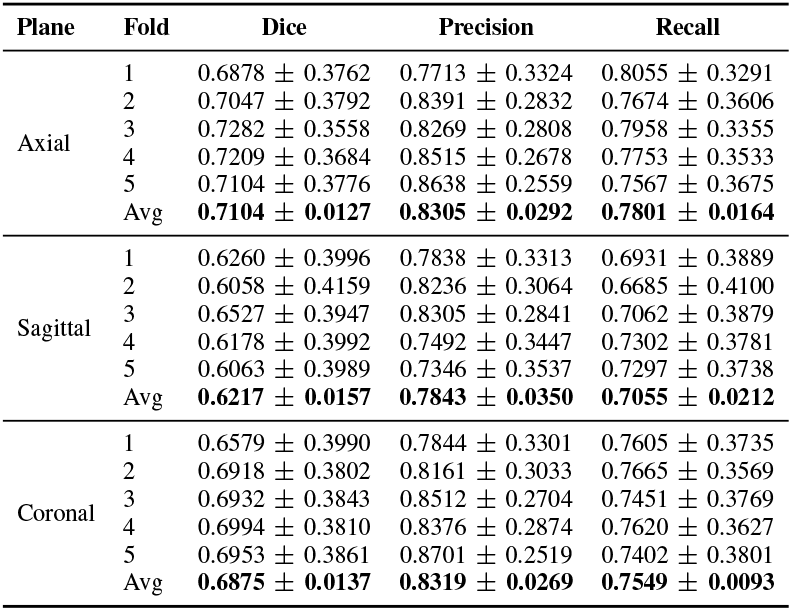
Five-Fold Performance of the Proposed Model on Axial, Sagittal, and Coronal Planes.

#### 2) Model Comparison: Evaluation Metrics

The proposed model (RAX-Net) was compared with representative methods (X-Net, and CLCI) under identical data splits and metrics. Table V reports the mean performance across folds per plane. RAX-Net achieved higher Dice and recall while maintaining competitive precision.

**TABLE V.** Comparison of RAX-Net with Representative Methods.

| Plane | Model | Dice | Precision | Recall | F1 |
| --- | --- | --- | --- | --- | --- |
| Axial | X-Net | 0.6606 | **0.9506** | 0.6590 | 0.7796 |
|  | CLCI | 0.5904 | 0.9760 | 0.5864 | 0.7310 |
|  | RAX | **0.7104** | 0.8305 | **0.7801** | **0.8045** |
| Sagittal | X-Net | 0.6103 | **0.9216** | 0.6165 | 0.7375 |
|  | CLCI | 0.5830 | 0.9000 | 0.6002 | 0.7200 |
|  | RAX | **0.6217** | 0.7843 | **0.7055** | **0.7429** |
| Coronal | X-Net | 0.6439 | **0.9585** | 0.6382 | 0.7666 |
|  | CLCI | 0.5721 | 0.6928 | 0.5774 | 0.6298 |
|  | RAX | **0.6875** | 0.8319 | **0.7549** | **0.7913** |

#### 3) Model Comparison: Computational Cost

Training and inference efficiency, as well as model size, are reported in Table VI. Across planes, RAX-Net required markedly fewer parameters and achieved lower training and prediction times.

**TABLE VI.** Average Training Time, Per-Slice Prediction Time, and Trainable Parameters.

| Plane | Model | Training Time (h) | Prediction Time (s) | Parameters |
| --- | --- | --- | --- | --- |
| Axial | X-Net | 8.98 | 0.0520 | 15,072,332 |
|  | CLCI | 3.91 | 0.0590 | 36,813,025 |
|  | RAX | **1.02** | **0.0418** | **4,829,665** |
| Sagittal | X-Net | 11.76 | 0.0540 | 15,072,332 |
|  | CLCI | 8.94 | 0.0620 | 36,813,025 |
|  | RAX | **1.75** | **0.0417** | **4,829,665** |
| Coronal | X-Net | 9.50 | 0.0520 | 15,072,332 |
|  | CLCI | 2.96 | 0.0980 | 36,813,025 |
|  | RAX | **1.31** | **0.0407** | **4,829,665** |

### C. Aggregation-Function Evaluation

#### 1) Per-Plane Evaluation on the Aggregation Set

The aggregation function was evaluated on 112 held-out cases. For each case, the best-performing model per plane (from cross-validation) generated plane-wise predictions; these were aggregated to produce final masks. Table VII shows that aggregation consistently improved Dice, precision, and recall across planes. An example for the axial plane (pre- and post-aggregation) is shown in Fig. 7.

**TABLE VII.** Per-Plane Performance Before and After Aggregation on 112 Cases.

| Plane | Aggregation | Dice | Precision | Recall |
| --- | --- | --- | --- | --- |
| Axial | No | 0.8016 | 0.8612 | 0.8748 |
|  | Yes | **0.8451** | **0.9019** | **0.8807** |
| Sagittal | No | 0.8231 | 0.8830 | 0.8951 |
|  | Yes | **0.8753** | **0.9331** | **0.9022** |
| Coronal | No | 0.8184 | 0.8984 | 0.8712 |
|  | Yes | **0.8633** | **0.9156** | **0.8987** |

**Fig. 7.**
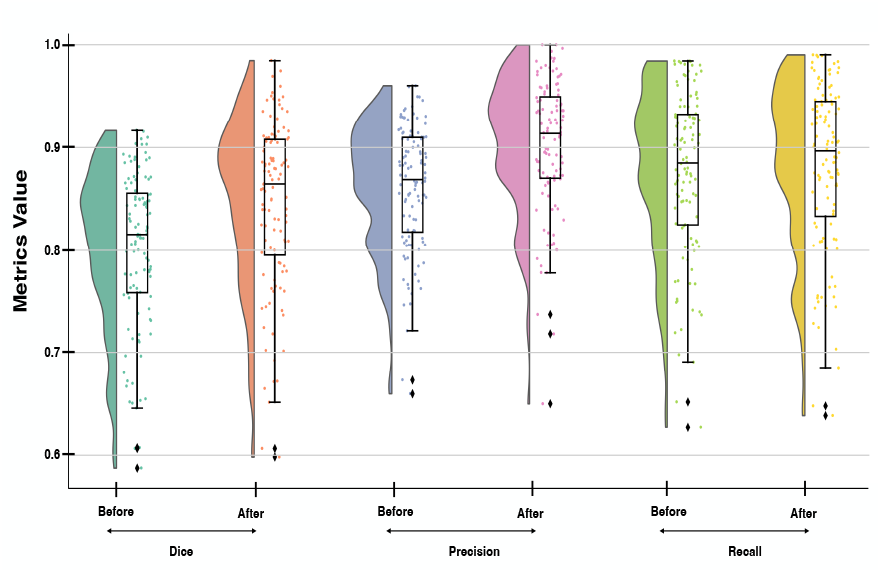
Axial-plane evaluation metrics before and after aggregation.

#### 2) Subject-Level 3D Evaluation

Aggregated plane-wise predictions were reconstructed into 3D masks and evaluated against the reference masks across the 112 cases. Figure 8 illustrates a representative 3D result, and Fig. 9 reports rain-cloud plots for Dice, precision, and recall over the evaluation set. Summary statistics are provided in Table VIII.

**Fig. 8.**
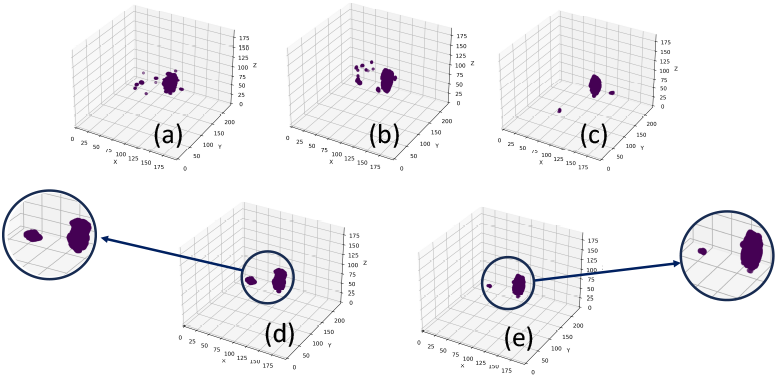
Representative 3D visualization: (a-c) planes (Axial, Sagittal, and Coronal), (d) ground-truth mask, and (e) final segmentation after aggregation.

**Fig. 9.**
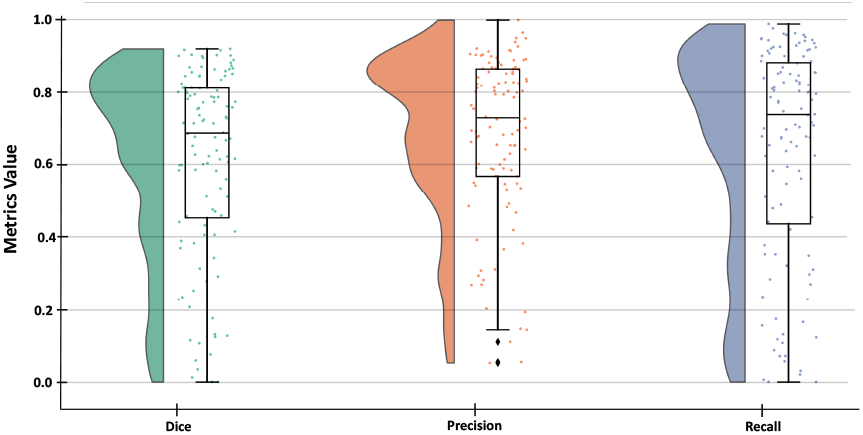
Raincloud plots of Dice, precision, and recall across 112 subjects.

**TABLE VIII.** Subject-Level Results on 112 3D MRI Cases (Aggregated Predictions)

| Dice | Precision | Recall | Volume Difference |
| --- | --- | --- | --- |
| $0.6126 \pm 0.2577$ | $0.6833 \pm 0.2281$ | $0.6385 \pm 0.2963$ | $6486 \pm 9686$ |

Note: volume difference ( $\text{mm}^3$ ).

## VI. Ablation Study

To assess the contribution of residual connections, the model was trained and evaluated on the sagittal split (fold 3) with and without residual connections. Results are reported in Table IX. Incorporating residual connections increased Dice and F1 while trading a small amount of precision for higher recall, indicating improved sensitivity and overall overlap.

**TABLE IX.** Effect of Residual Connections on Segmentation Performance (Sagittal, Fold 3)

|  | Dice | Precision | Recall | F1 |
| --- | --- | --- | --- | --- |
| With residual | 0.6260 | 0.7838 | 0.6931 | 0.7356 |
| Without residual | 0.5951 | 0.8070 | 0.6403 | 0.7147 |

### Interpretation

Residual connections yielded ΔDice = +0.0309 and ΔF1 = +0.0209, primarily via increased recall (+0.0528) with a modest reduction in precision (− 0.0232). This pattern suggests that residual pathways facilitate recovery of lesion voxels that are otherwise missed, improving boundary recall and overlap.

## VII. Conclusion

RAX-Net, a residual attention Xception based architecture for ischemic stroke lesion segmentation in T1-weighted MRI, was presented. By combining separable convolutions, residual connections, and up-sampling residual attention blocks, the approach attains a favorable balance between computational efficiency and segmentation accuracy. A focal–Tversky loss variant further improved robustness to class imbalance and enhanced sensitivity to small or low-contrast lesion boundaries.

Extensive experiments on the ATLAS v2.0 dataset demonstrated performance gains over representative state-of-the-art methods in terms of Dice, recall, and F1 score, alongside markedly fewer trainable parameters and reduced training and inference times. The multi-plane decomposition with voxel-wise aggregation effectively leveraged complementary anatomical views to improve 3D segmentation quality.

These findings indicate the potential of RAX-Net as a practical tool for stroke lesion analysis, enabling more consistent and scalable segmentation than manual annotation. Future work will extend the framework to multimodal MRI and investigate integration into clinical workflows, including longitudinal monitoring and prognosis prediction.

## Data Availability

All data produced in the present work are contained in the manuscript

## Notes

### Competing Interest Statement

The authors have declared no competing interest.

### Funding Statement

This study did not receive any funding

### Author Declarations

ATLAS 2 Source data:http://fcon_1000.projects.nitrc.org/indi/retro/atlas.html Article: https://www.medrxiv.org/content/10.1101/2021.12.09.21267554v1

## References

[1] S. S. Martin et al., “2025 Heart Disease and Stroke Statistics: A Report of US and Global Data From the American Heart Association,” Circulation, vol. 151, no. 8, Feb. 2025, doi: 10.1161/CIR.0000000000001303.

[2] R. L. Sacco et al., “An Updated Definition of Stroke for the 21st Century,” Stroke, vol. 44, no. 7, pp. 2064–2089, Jul. 2013, doi: 10.1161/STR.0b013e318296aeca.

[3] O. Maier et al., “ISLES 2015 – a public evaluation benchmark for ischemic stroke lesion segmentation from multispectral MRI,” Med. Image Anal., vol. 35, pp. 250–269, Jan. 2017, doi: 10.1016/j.media.2016.07.009.

[4] J. M. Biesbroek, H. J. Kuijf, N. A. Weaver, L. Zhao, M. Duering, and G. J. Biessels, “Brain infarct segmentation and registration on MRI or CT for lesion-symptom mapping,” J. Vis. Exp., no. 151, Sep. 2019, doi: 10.3791/59653.

[5] D. Smajlović and O. Sinanović, “Sensitivity of the neuroimaging techniques in ischemic stroke,” Med. Arh., vol. 58, no. 5, pp. 282–284, 2004.

[6] A. B. Neumann et al., “Interrater agreement for final infarct MRI lesion delineation,” Stroke, vol. 40, no. 12, pp. 3768–3771, Dec. 2009, doi: 10.1161/STROKEAHA.108.545368.

[7] L. Tomasetti, L. J. Hollesli, K. Engan, K. D. Kurz, M. W. Kurz, and M. Khanmohammadi, “Machine learning algorithms versus thresholding to segment ischemic regions in patients with acute ischemic stroke,” IEEE J. Biomed. Health Inform., vol. 26, no. 2, pp. 660–672, Feb. 2022, doi: 10.1109/JBHI.2021.3097591.

[8] K. L. Ito, H. Kim, and S. Liew, “A comparison of automated lesion segmentation approaches for chronic stroke T1-weighted MRI data,” Hum. Brain Mapp., vol. 40, no. 16, pp. 4669–4685, Nov. 2019, doi: 10.1002/hbm.24729.

[9] W. Qiu et al., “Machine learning for detecting early infarction in acute stroke with non-contrast-enhanced CT,” Radiology, vol. 294, no. 3, pp. 638–644, Mar. 2020, doi: 10.1148/radiol.2020191193.

[10] A. S. Kasasbeh, S. Christensen, M. W. Parsons, B. Campbell, G. W. Albers, and M. G. Lansberg, “Artificial neural network computer tomography perfusion prediction of ischemic core,” Stroke, vol. 50, no. 6, pp. 1578–1581, Jun. 2019, doi: 10.1161/STROKEAHA.118.022649.

[11] W. Qiu et al., “Automated prediction of ischemic brain tissue fate from multiphase computed tomographic angiography in patients with acute ischemic stroke using machine learning,” J. Stroke, vol. 23, no. 2, pp. 234–243, May 2021, doi:10.5853/jos.2020.05064.

[12] S.-L. Liew et al., “A large, curated, open-source stroke neuroimaging dataset to improve lesion segmentation algorithms,” Sci. Data, vol. 9, no. 1, p. 320, Jun. 2022, doi: 10.1038/s41597-022-01401-7.

[13] Y. Zhang, S. Liu, C. Li, and J. Wang, “Application of deep learning method on ischemic stroke lesion segmentation,” J. Shanghai Jiaotong Univ. (Sci.), vol. 27, no. 1, pp. 99–111, Feb. 2022, doi: 10.1007/s12204-021-2273-9.

[14] O. Ronneberger, P. Fischer, and T. Brox, “U-net: Convolutional networks for biomedical image segmentation,” in Med. Image Comput. Comput.-Assist. Intervent. (MICCAI), Cham, Switzerland: Springer, 2015, pp. 234–241, doi: 10.1007/978-3-319-24574-428.

[15] L. Liu, F.-X. Wu, and J. Wang, “Efficient multi-kernel DCNN with pixel dropout for stroke MRI segmentation,” Neurocomputing, vol. 350, pp. 117–127, Jul. 2019, doi: 10.1016/j.neucom.2019.03.049.

[16] R. Sathish, R. Rajan, A. Vupputuri, N. Ghosh, and D. Sheet, “Adversarially trained convolutional neural networks for semantic segmentation of ischaemic stroke lesion using multisequence magnetic resonance imaging,” in Proc. Annu. Int. Conf. IEEE Eng. Med. Biol. Soc. (EMBC), Jul. 2019, pp. 1010–1013, doi: 10.1109/EMBC.2019.8857527.

[17] A. Kumar et al., “CSNet: A new DeepNet framework for ischemic stroke lesion segmentation,” Comput. Methods Programs Biomed., vol. 193, p. 105524, Sep. 2020, doi: 10.1016/j.cmpb.2020.105524.

[18] A. Clérigues, S. Valverde, J. Bernal, J. Freixenet, A. Oliver, and X. Lladó, “Acute and sub-acute stroke lesion segmentation from multimodal MRI,” Comput. Methods Programs Biomed., vol. 194, p. 105521, Oct. 2020, doi: 10.1016/j.cmpb.2020.105521.

[19] L. Liu, S. Chen, F. Zhang, F.-X. Wu, Y. Pan, and J. Wang, “Deep convolutional neural network for automatically segmenting acute ischemic stroke lesion in multi-modality MRI,” Neural Comput. Appl., vol. 32, no. 11, pp. 6545–6558, Jun. 2020, doi: 10.1007/s00521-019-04096-x.

[20] K. Kamnitsas et al., “Efficient multi-scale 3D CNN with fully connected CRF for accurate brain lesion segmentation,” Med. Image Anal., vol. 36, pp. 61–78, Feb. 2017, doi: 10.1016/j.media.2016.10.004.

[21] L. Zhang et al., “Ischemic stroke lesion segmentation using multi-plane information fusion,” IEEE Access, vol. 8, pp. 45715–45725, 2020, doi: 10.1109/ACCESS.2020.2977415.

[22] L. Liu, L. Kurgan, F.-X. Wu, and J. Wang, “Attention convolutional neural network for accurate segmentation and quantification of lesions in ischemic stroke disease,” Med. Image Anal., vol. 65, p. 101791, Oct. 2020, doi: 10.1016/j.media.2020.101791.

[23] Y. Zhou, W. Huang, P. Dong, Y. Xia, and S. Wang, “D-UNet: A dimension-fusion U-shape network for chronic stroke lesion segmentation,” IEEE/ACM Trans. Comput. Biol. Bioinf., vol. 18, no. 3, pp. 940–950, 2021, doi: 10.1109/TCBB.2019.2939522.

[24] X. Liu et al., “MSDF-net: Multi-scale deep fusion network for stroke lesion segmentation,” IEEE Access, vol. 7, pp. 178486–178495, 2019, doi: 10.1109/ACCESS.2019.2958384.

[25] K. Qi et al., “X-net: Brain stroke lesion segmentation based on depthwise separable convolution and long-range dependencies,” in Med. Image Comput. Comput.-Assist. Intervent. (MICCAI), Cham, Switzerland: Springer, 2019, pp. 247–255, doi: 10.1007/978-3-030-32248-928.

[26] H. Yang et al., “CLCI-net: Cross-level fusion and context inference networks for lesion segmentation of chronic stroke,” in Med. Image Comput. Comput.-Assist. Intervent. (MICCAI), Cham, Switzerland: Springer, 2019, pp. 266–274, doi: 10.1007/978-3-030-32248-930.

[27] H. Hui, X. Zhang, F. Li, X. Mei, and Y. Guo, “A partitioning-stacking prediction fusion network based on an improved attention U-net for stroke lesion segmentation,” IEEE Access, vol. 8, pp. 47419–47432, 2020, doi: 10.1109/ACCESS.2020.2977946.

[28] N. Tomita, S. Jiang, M. E. Maeder, and S. Hassanpour, “Automatic post-stroke lesion segmentation on MR images using 3D residual convolutional neural network,” NeuroImage Clin., vol. 27, p. 102276, 2020, doi: 10.1016/j.nicl.2020.102276.

[29] J. Shore and R. Johnson, “Axiomatic derivation of the principle of maximum entropy and the principle of minimum cross-entropy,” IEEE Trans. Inf. Theory, vol. 26, no. 1, pp. 26–37, Jan. 1980, doi: 10.1109/TIT.1980.1056144.

[30] F. Milletari, N. Navab, and S.-A. Ahmadi, “V-net: Fully convolutional neural networks for volumetric medical image segmentation,” in Proc. Int. Conf. 3D Vision (3DV), Stanford, CA, USA, Oct. 2016, pp. 565–571, doi: 10.1109/3DV.2016.79.

[31] T.-Y. Lin, P. Goyal, R. Girshick, K. He, and P. Dollar, “Focal loss for dense object detection,” in Proc. IEEE Int. Conf. Comput. Vis. (ICCV), Venice, Italy, Oct. 2017, pp. 2980–2988.

[32] S. S. M. Salehi, D. Erdogmus, and A. Gholipour, “Tversky loss function for image segmentation using 3D fully convolutional deep networks,” in Med. Image Comput. Comput.-Assist. Intervent. (MICCAI), Cham, Switzerland: Springer, 2017, pp. 379–387, doi: 10.1007/978-3-319-67389-944.

